# The impact of COVID-19 pandemic on influenza transmission: molecular and epidemiological evidence

**DOI:** 10.1101/2021.06.08.21258434

**Authors:** Leon King Tran, Dai-Wei Huang, Nien-Kung Li, Lucy M. Li, Julia A. Palacios, Hsiao-Han Chang

## Abstract

To quantify the impact of COVID-19-related control measures on the spread of human influenza virus, we analyzed case numbers, viral molecular sequences, personal behavior data, and policy stringency data from various countries, and found consistent evidence of decrease in influenza incidence after the emergence of COVID-19.

**Article Summary:** We quantify a noticeable decrease in H1N1 and H3N2 cases and genetic diversity in selected countries since the onset of the COVID-19 pandemic.

## Introduction

The emergence and spread of COVID-19 in 2020 led to a number of large-scale public health measures to limit international travel, reduce gatherings, and increase mask-wearing. While these preventative measures were implemented to curtail the COVID-19 pandemic, they seem to have also impacted the spread of other respiratory illnesses. There have been several reports on the decrease in case numbers during 2019-2020 influenza season in the northern hemisphere [6], and the lack of a 2020 influenza season in the southern hemisphere [8]. Here, we quantify the impact of the COVID-19 pandemic on the spread of influenza in terms of incidence and viral molecular diversity [1].

## Method

### Case count data

We analyzed weekly case count data of influenza available in FluNet [10] from various regions during the 2010-2020 influenza seasons. We defined *T*_*S*_ and *T*_*E*_ as the weeks during which the estimated number of cases reached 10% and 90% of the total case numbers in each influenza season, respectively. Since the influenza outbreaks for most regions started before the COVID-19 outbreak, we compared *T*_*E*_ and durations of influenza seasons pre- and post-COVID-19 pandemic. We defined the duration of an influenza season by the difference between *T*_*S*_ and *T*_*E*_, and standardized the duration in the 2019-2020 season by the average and standard deviation of the duration from previous 9 seasons.

### Viral molecular sequences

We analyzed the HA segment of human influenza A H1N1 and H3N2 sequences available in the GISAID EpiFlu database on November 1^st^ 2020. The collection dates of the sequences ranged from January 2016 to December 2020. We used BEAST [3] to estimate the effective population size (N_e_) from 2016 to 2020 for each location. The numbers of sequences analyzed are indicated in Appendix Table 1.

## Results and Discussion

To examine the indirect impact of COVID-19 on influenza dynamics, we compared the 2019-2020 influenza season with previous 9 seasons in 21 locations across 5 continents (Table 1). We found that for all locations in Asia, the outbreak in 2019-2020 both ended earlier and lasted for a shorter duration than previous years. In the Americas, Europe, and Africa, 12 locations out of 15 showed an earlier end of the flu season in the 2019-2020 season than previous years; the rest remained similar. For locations where influenza seasons usually end later in the year, such as Brazil, Guatemala, and South Africa, the difference in duration between the 2019-2020 season and previous seasons was larger than other locations. The flu season in the Southern Hemisphere, which usually starts much later in the year, disappeared in several countries in 2020 [6,8].

In addition to case count data, we analyzed molecular data to evidence our findings. For each location and each influenza type, we calculated the within-location genetic diversity, Watterson’s θ [9], for the first half of 2019 and for the first of 2020 (Appendix Table 2). We found that for 10 out of 11 locations we analyzed, θ decreased from 2019 to 2020 for H1N1, and 9 out of 11 locations for H3N2. On the other hand, we calculated between-region genetic diversity for each pair of regions once in 2019 and again in 2020; 9 out of 11 locations for H1N1 and 6 out of 11 locations for H3N2 had their between-region genetic diversity increase from 2019 to 2020 in at least 50% of pairs for which the location was involved, reflecting reduced travel between regions in 2020 (Appendix Table 3).

**Table 1.**
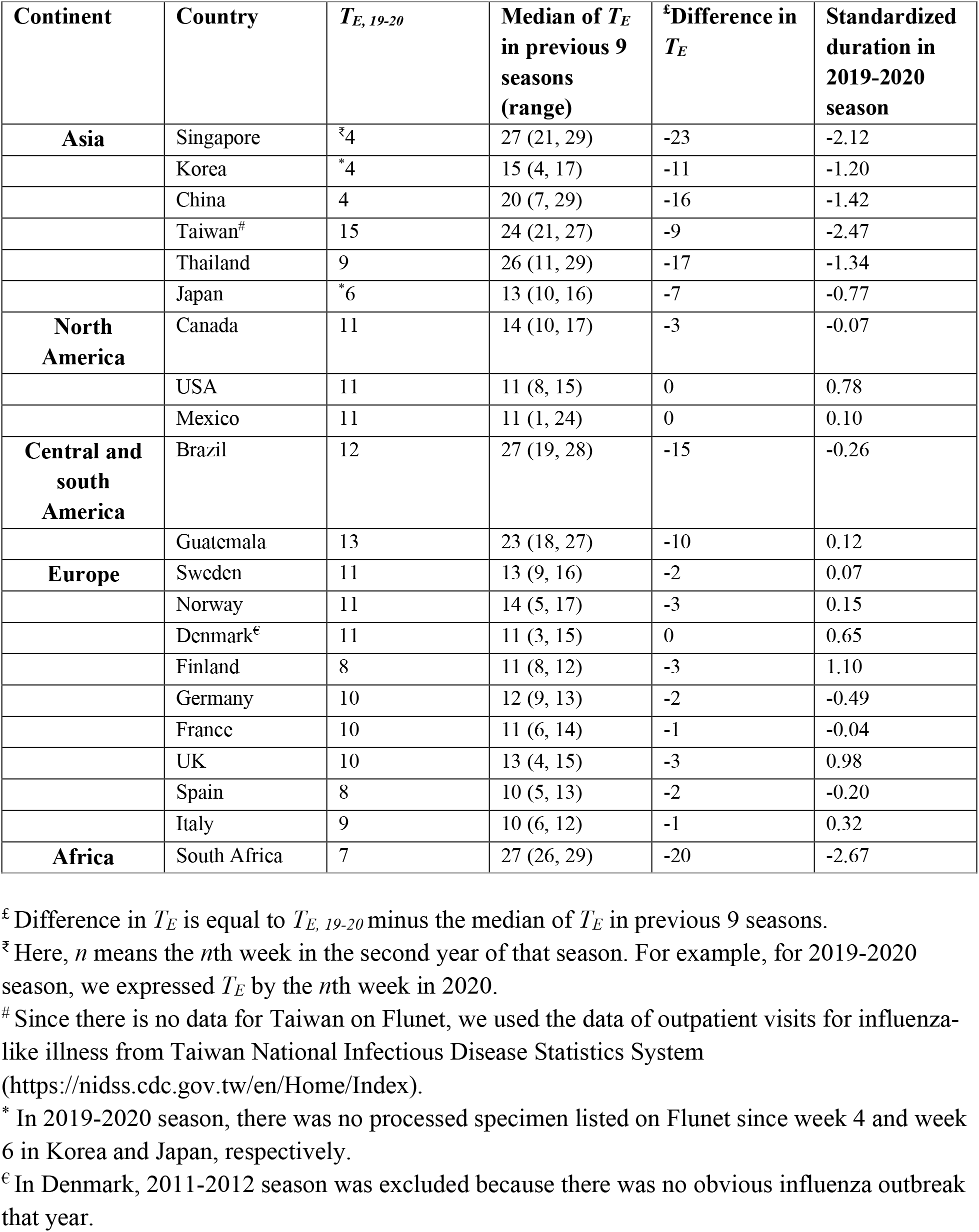
Comparison of influenza outbreaks before and after the emergence of COVID-19.

While Watterson’s θ measures overall viral diversity, the effective population size (N_e_) quantifies genetic diversity over time [4]. We estimated N_e_ for H1N1 in 11 countries and found a decrease in N_e_ in 9 countries, including Italy and Taiwan (Figure 1; Appendix Figure 1; Appendix Table 1). For H3N2, we analyzed 5 countries, and found a decrease in N_e_ in South Africa and Taiwan in 2020 (Figure 1; Appendix Figure 2; Appendix Table 1).

**Figure 1.**
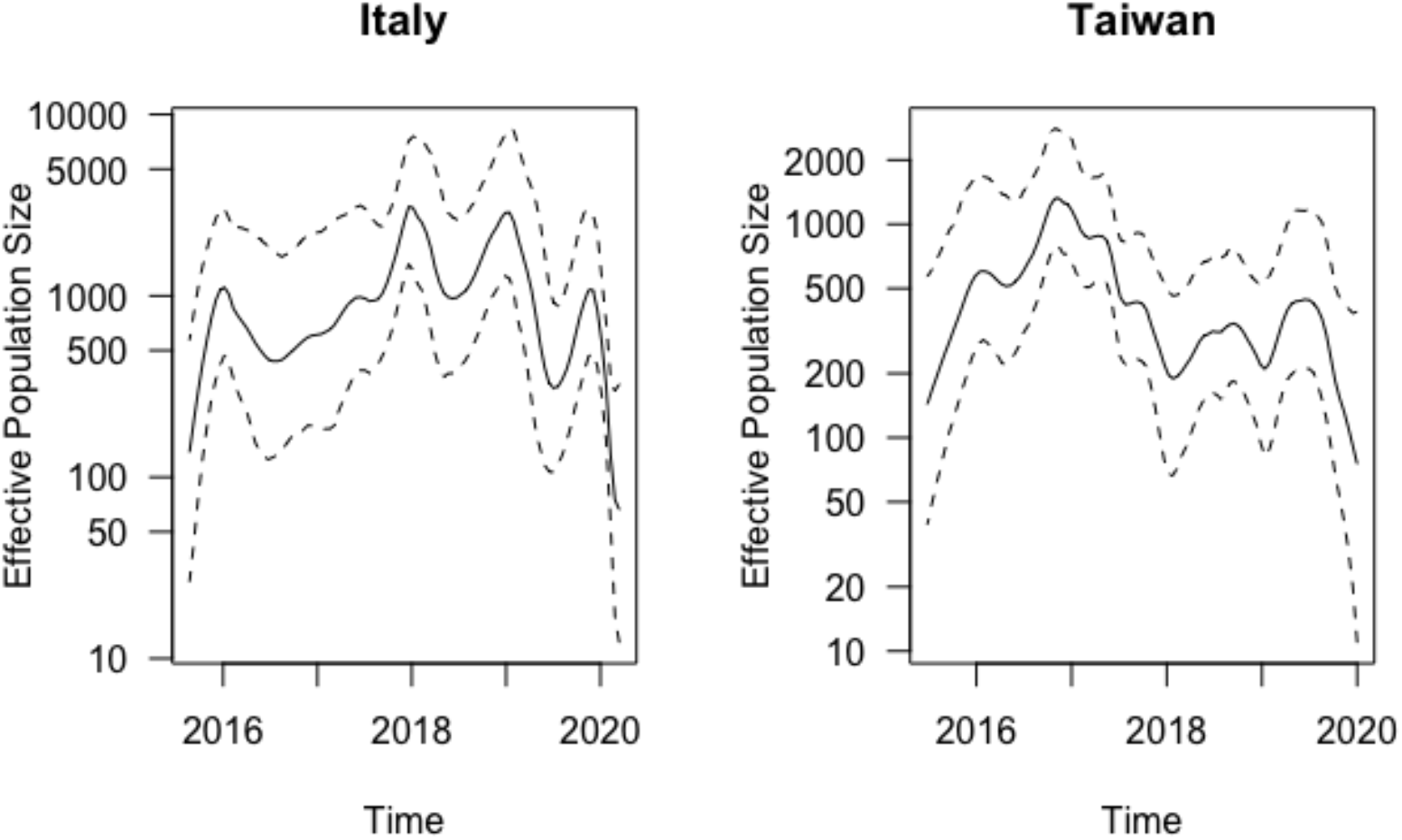
Effective Population Size Trajectory of H1N1 in Italy and H3N2 in Taiwan.

For personal measures and government policies against COVID-19, we also noticed that Asian countries tend to act earlier than countries in other continents, especially wearing masks (Appendix Table 4). Taken together, we observed earlier ends of flu seasons in Asia than in Europe and America, which could be explained by the earlier implementation of non-pharmaceutical interventions.

The decrease in influenza incidence after the emergence of COVID-19 was confirmed by both case count and molecular data, providing stronger support for a genuine decrease in influenza incidence, even when count data might be incomplete or imprecise due to potentially less flu surveillance in 2020.

## Data Availability

The flu sequences are found on GISAID. The accession numbers can be found at https://github.com/leonkt/accession-numbers.

https://www.gisaid.org/

## Acknowledgements

We thank GISAID and sequencing laboratories for making the influenza A viral molecular sequences available Specific laboratories and accession numbers are found at the end of the Appendix. HHC and DWH were supported by the Ministry of Science and Technology in Taiwan (MOST 110-2636-B-007-009); HHC was supported by the Yushan Scholar Program. The funders had no role in study design, data collection, data analysis, data interpretation, or writing of the report.

## First Author Biography

Leon King Tran is an undergraduate student in Mathematics and master’s student in Statistics at Stanford University and is interested in Biostatistics and Stochastic Processes.

Dai-Wei Huang is an undergraduate student in the Interdisciplinary Program of Life Science, National Tsing Hua University, Hsinchu, Taiwan and is currently doing research related to epidemiology and biostatistics.

## Appendix

## Methods

### Personal measures and governmental policies against COVID-19

We analyzed two personal measures taken against the spread of COVID-19 from YouGov [7]– wearing masks when in public places and improving personal hygiene, from February 21, 2020 to December 17, 2020. For governmental responses to COVID-19, we used the data from OxCGRT, the Oxford COVID-19 Government Response Tracker [5], from January 1, 2020 to May 31, 2020.

### Molecular Epidemiology

We estimated a mutation rate of H3N2 for New York State of 4 x 10^−3^ per base per year and 3.6 x 10^−3^ for H1N1. We then fixed the corresponding global mutation rates for the rest of the locations. We assumed the HKY prior mutation model with empirical frequencies and Gamma+Invariant site heterogeneity with 4 Gamma categories and 3 partitions into codon positions and assumed the Bayesian Skyride Coalescent prior on effective population size trajectories. We examined convergence by the likelihood trace plot and the effective sample size for each location and type. To estimate N_e_, we divided the output (N_e_ * G_t_) by the generation time (G_t_) for H1N1 (2.3 days) and for H3N2 (3.1 days) [2]. We ran the MCMC for 10,000,000 iterations, thinning every 1000 iterations and with 10% of burnin. We removed any regions without convergence.

The within-region diversities were measured by Watterson’s θ. The between-region diversities were measured as follows:

Between-region diversity between region 1 and region 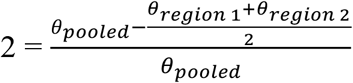, where *θ*_*pooled*_ was Watterson’s θ when combining two regions together.

**Appendix Table 1.** Effective sample sizes of root age and number of sequences per country for influenza types H1N1 and H3N2. These effective sample size numbers were obtained with Tracer [3]. Effective sample sizes with N/A indicate lack of convergence and were omitted from the analysis.

| Country | Genotype | Effective Sample Size | Sequences |
| --- | --- | --- | --- |
| **Australia** | H1N1 | 7 | 749 |
| **Bangladesh** | H1N1 | 15 | 407 |
| **Brazil** | H1N1 | N/A | 722 |
| **California, USA** | H1N1 | N/A | 514 |
| **Chile** | H1N1 | 9 | 741 |
| **China** | H1N1 | 6 | 674 |
| **England, UK** | H1N1 | N/A | 939 |
| **France** | H1N1 | 5 | 905 |
| **Guatemala** | H1N1 | 1546 | 126 |
| **Italy** | H1N1 | 145 | 250 |
| **Japan** | H1N1 | 19 | 782 |
| **New York, USA** | H1N1 | 2044 | 186 |
| **South Africa** | H1N1 | 3553 | 180 |
| **Taiwan** | H1N1 | 3392 | 115 |
| **Australia** | H3N2 | N/A | 778 |
| **Bangladesh** | H3N2 | 18 | 503 |
| **Brazil** | H3N2 | N/A | 774 |
| **California, USA** | H3N2 | N/A | 820 |
| **Chile** | H3N2 | N/A | 625 |
| **China** | H3N2 | N/A | 958 |
| **England, UK** | H3N2 | N/A | 997 |
| **France** | H3N2 | N/A | 727 |
| **Guatemala** | H3N2 | 2407 | 145 |
| **Italy** | H3N2 | N/A | 513 |
| **Japan** | H3N2 | 4 | 795 |
| **New York, USA** | H3N2 | N/A | 625 |
| **South Africa** | H3N2 | 220 | 223 |
| **Taiwan** | H3N2 | 550 | 331 |

**Appendix Table 2.** Within-region genetic diversity during the first half of 2019 compared to the first half of 2020, for influenza types H1N1 and H3N2. Diversity was measured by Watterson’s θ. The decrease column indicates whether genetic diversity decreased from the first half of 2019 to the first half of 2020. Sequences with more than 5% gaps were removed from the analysis. N/A indicates a lack of data.

| Country/Region | Genotype | 2019 | 2020 | Decrease |
| --- | --- | --- | --- | --- |
| Australia | H1N1 | 2.77E-02 | 2.20E-02 | Yes |
| Bangladesh | H1N1 | N/A | N/A | N/A |
| Brazil | H1N1 | 2.12E-02 | 1.38E-02 | Yes |
| California, USA | H1N1 | 2.55E-02 | 1.63E-02 | Yes |
| Chile | H1N1 | 2.22E-02 | 9.46E-03 | Yes |
| China | H1N1 | 3.46E-02 | 1.75E-02 | Yes |
| England, UK | H1N1 | 2.93E-02 | 2.03E-02 | Yes |
| France | H1N1 | 2.51E-02 | 2.08E-02 | Yes |
| Italy | H1N1 | 2.03E-02 | 1.28E-02 | Yes |
| Japan | H1N1 | 2.64E-02 | 1.88E-02 | Yes |
| New York, USA | H1N1 | 1.94E-02 | 1.68E-02 | Yes |
| South Africa | H1N1 | 6.44E-03 | 8.76E-03 | No |
| Taiwan | H1N1 | N/A | N/A | N/A |
| Australia | H3N2 | 2.97E-02 | 2.32E-02 | Yes |
| Bangladesh | H3N2 | N/A | N/A | N/A |
| Brazil | H3N2 | 2.65E-02 | 2.68E-02 | No |
| California, USA | H3N2 | 2.35E-02 | 4.58E-02 | No |
| Chile | H3N2 | 2.11E-02 | 1.35E-02 | Yes |
| China | H3N2 | 5.99E-02 | 1.45E-02 | Yes |
| England, UK | H3N2 | 2.77E-02 | 2.15E-02 | Yes |
| France | H3N2 | 3.28E-02 | 2.25E-02 | Yes |
| Italy | H3N2 | 2.66E-02 | 1.87E-02 | Yes |
| Japan | H3N2 | 3.02E-02 | 1.16E-02 | Yes |
| New York, USA | H3N2 | 1.70E-02 | 8.22E-03 | Yes |
| South Africa | H3N2 | 1.86E-02 | 3.21E-03 | Yes |
| Taiwan | H3N2 | N/A | N/A | N/A |

**Appendix Table 3.**
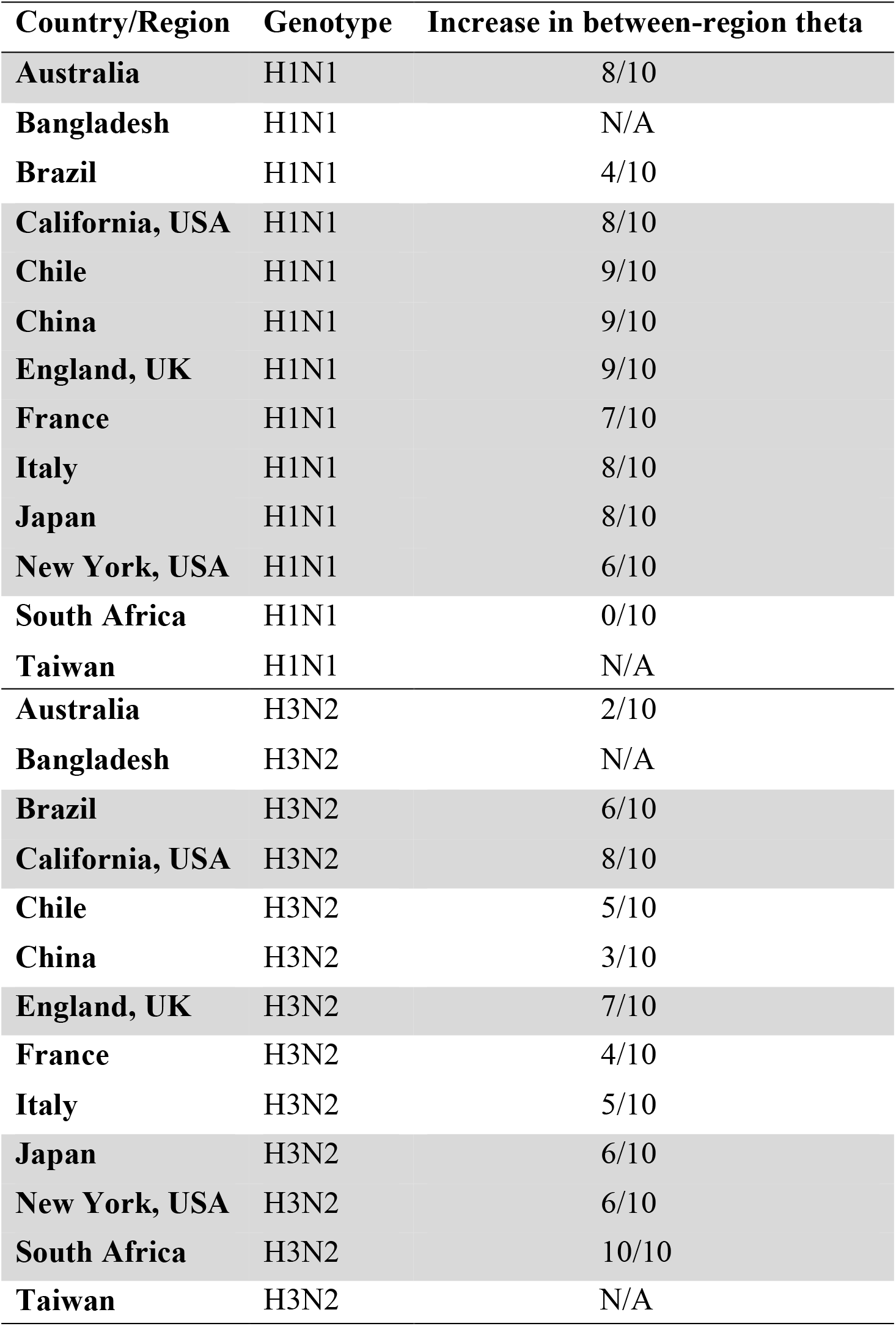
Between-region diversity during the first half of 2019 compared to the first half of 2020, for influenza types H1N1 and H3N2. For each pair of regions, we calculated the between-region diversity in the first half of 2019 and in the first half of 2020. The third column for each region denotes the number of pairs involving that region with an increase in between-region diversity from 2019 to 2020, divided by the total number of pairwise comparisons. Regions where this fraction is above 0.5 are marked in grey. N/A indicates that there was not enough data to calculate diversity.

| Country/Region | Genotype | Increase in between-region theta |
| --- | --- | --- |
| **Australia** | H1N1 | 8/10 |
| **Bangladesh** | H1N1 | N/A |
| **Brazil** | H1N1 | 4/10 |
| **California, USA** | H1N1 | 8/10 |
| **Chile** | H1N1 | 9/10 |
| **China** | H1N1 | 9/10 |
| **England, UK** | H1N1 | 9/10 |
| **France** | H1N1 | 7/10 |
| **Italy** | H1N1 | 8/10 |
| **Japan** | H1N1 | 8/10 |
| **New York, USA** | H1N1 | 6/10 |
| **South Africa** | H1N1 | 0/10 |
| **Taiwan** | H1N1 | N/A |
| **Australia** | H3N2 | 2/10 |
| **Bangladesh** | H3N2 | N/A |
| **Brazil** | H3N2 | 6/10 |
| **California, USA** | H3N2 | 8/10 |
| **Chile** | H3N2 | 5/10 |
| **China** | H3N2 | 3/10 |
| **England, UK** | H3N2 | 7/10 |
| **France** | H3N2 | 4/10 |
| **Italy** | H3N2 | 5/10 |
| **Japan** | H3N2 | 6/10 |
| **New York, USA** | H3N2 | 6/10 |
| **South Africa** | H3N2 | 10/10 |
| **Taiwan** | H3N2 | N/A |

**Appendix Table 4.**
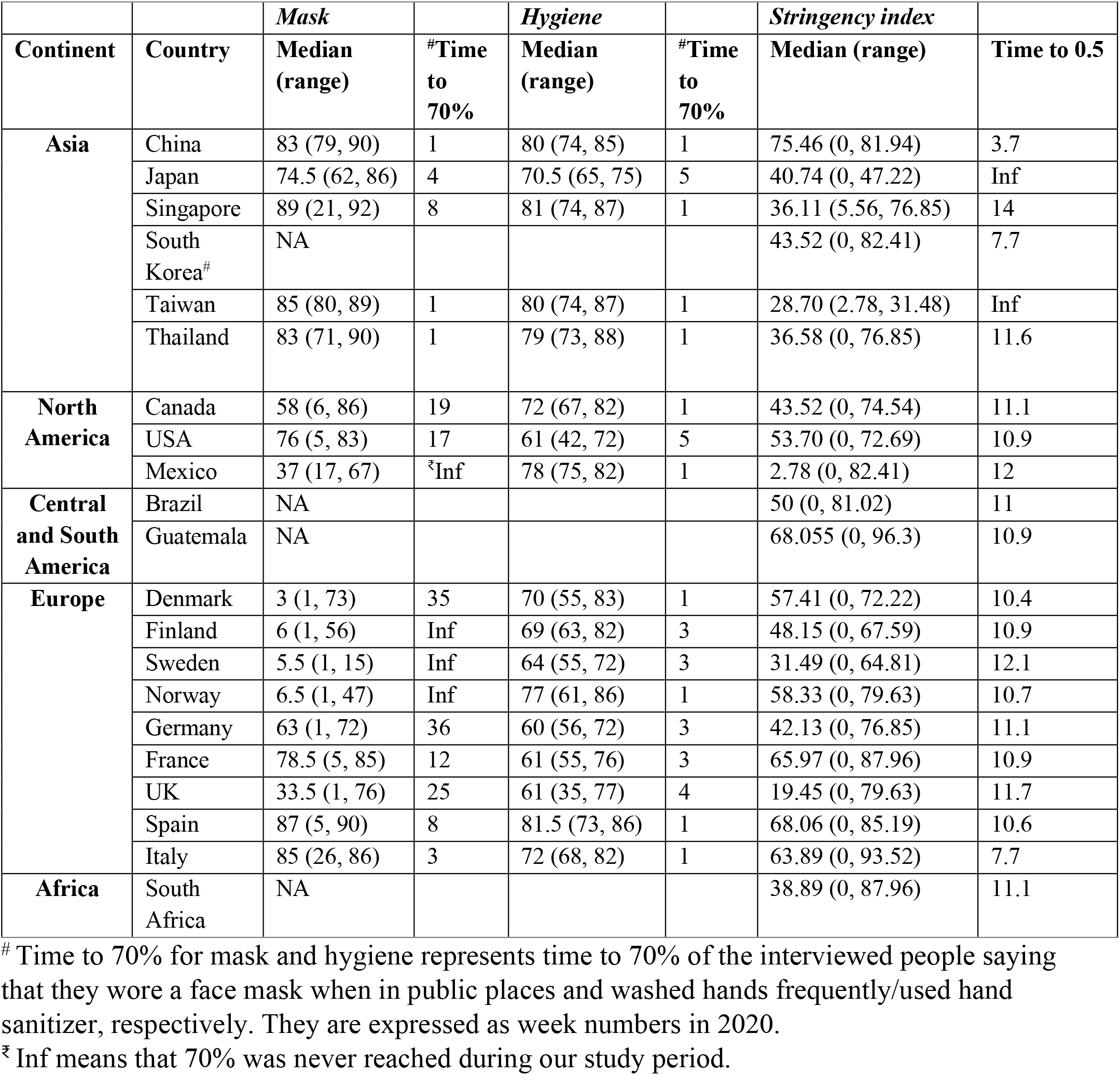
Comparison of personal measures and governmental policies against COVID-19

|  |  | *Mask* |  | *Hygiene* |  | *Stringency index* |  |
| --- | --- | --- | --- | --- | --- | --- | --- |
| **Continent** | **Country** | **Median (range)** | **#Time to 70%** | **Median (range)** | **#Time to 70%** | **Median (range)** | **Time to 0.5** |
| **Asia** | China | 83 (79, 90) | 1 | 80 (74, 85) | 1 | 75.46 (0, 81.94) | 3.7 |
|  | Japan | 74.5 (62, 86) | 4 | 70.5 (65, 75) | 5 | 40.74 (0, 47.22) | Inf |
|  | Singapore | 89 (21, 92) | 8 | 81 (74, 87) | 1 | 36.11 (5.56, 76.85) | 14 |
|  | South Korea<sup>#</sup> | NA |  |  |  | 43.52 (0, 82.41) | 7.7 |
|  | Taiwan | 85 (80, 89) | 1 | 80 (74, 87) | 1 | 28.70 (2.78, 31.48) | Inf |
|  | Thailand | 83 (71, 90) | 1 | 79 (73, 88) | 1 | 36.58 (0, 76.85) | 11.6 |
| **North America** | Canada | 58 (6, 86) | 19 | 72 (67, 82) | 1 | 43.52 (0, 74.54) | 11.1 |
|  | USA | 76 (5, 83) | 17 | 61 (42, 72) | 5 | 53.70 (0, 72.69) | 10.9 |
|  | Mexico | 37 (17, 67) | <sup>‡</sup>Inf | 78 (75, 82) | 1 | 2.78 (0, 82.41) | 12 |
| **Central and South America** | Brazil | NA |  |  |  | 50 (0, 81.02) | 11 |
|  | Guatemala | NA |  |  |  | 68.055 (0, 96.3) | 10.9 |
| **Europe** | Denmark | 3 (1, 73) | 35 | 70 (55, 83) | 1 | 57.41 (0, 72.22) | 10.4 |
|  | Finland | 6 (1, 56) | Inf | 69 (63, 82) | 3 | 48.15 (0, 67.59) | 10.9 |
|  | Sweden | 5.5 (1, 15) | Inf | 64 (55, 72) | 3 | 31.49 (0, 64.81) | 12.1 |
|  | Norway | 6.5 (1, 47) | Inf | 77 (61, 86) | 1 | 58.33 (0, 79.63) | 10.7 |
|  | Germany | 63 (1, 72) | 36 | 60 (56, 72) | 3 | 42.13 (0, 76.85) | 11.1 |
|  | France | 78.5 (5, 85) | 12 | 61 (55, 76) | 3 | 65.97 (0, 87.96) | 10.9 |
|  | UK | 33.5 (1, 76) | 25 | 61 (35, 77) | 4 | 19.45 (0, 79.63) | 11.7 |
|  | Spain | 87 (5, 90) | 8 | 81.5 (73, 86) | 1 | 68.06 (0, 85.19) | 10.6 |
|  | Italy | 85 (26, 86) | 3 | 72 (68, 82) | 1 | 63.89 (0, 93.52) | 7.7 |
| **Africa** | South Africa | NA |  |  |  | 38.89 (0, 87.96) | 11.1 |

<sup>#</sup> Time to 70% for mask and hygiene represents time to 70% of the interviewed people saying that they wore a face mask when in public places and washed hands frequently/used hand sanitizer, respectively. They are expressed as week numbers in 2020.
<sup>‡</sup> Inf means that 70% was never reached during our study period.

**Appendix Figure 1.**
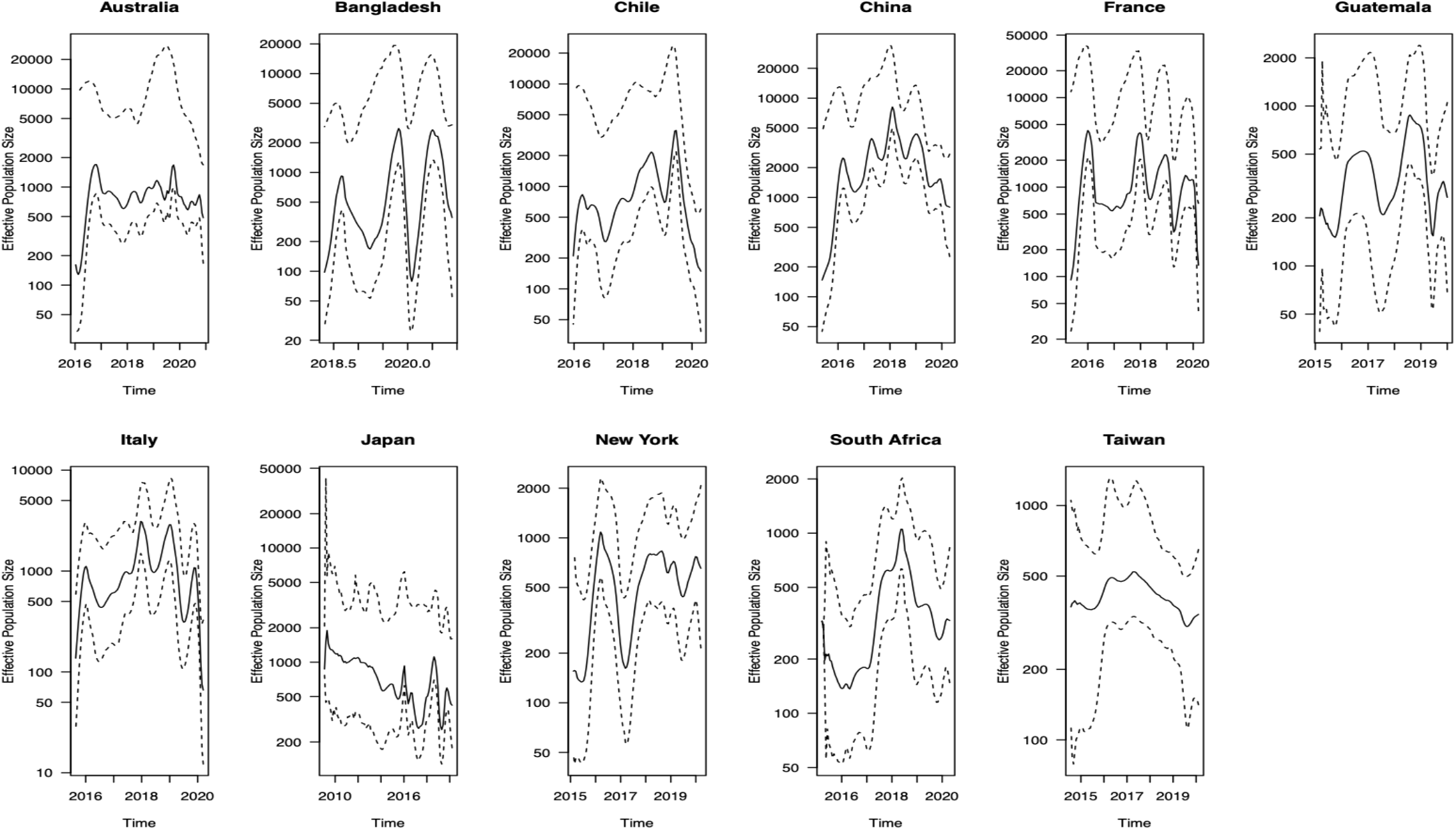
Estimated Effective Population Size Trajectory for H1N1 in selected regions.

**Appendix Figure 2.**
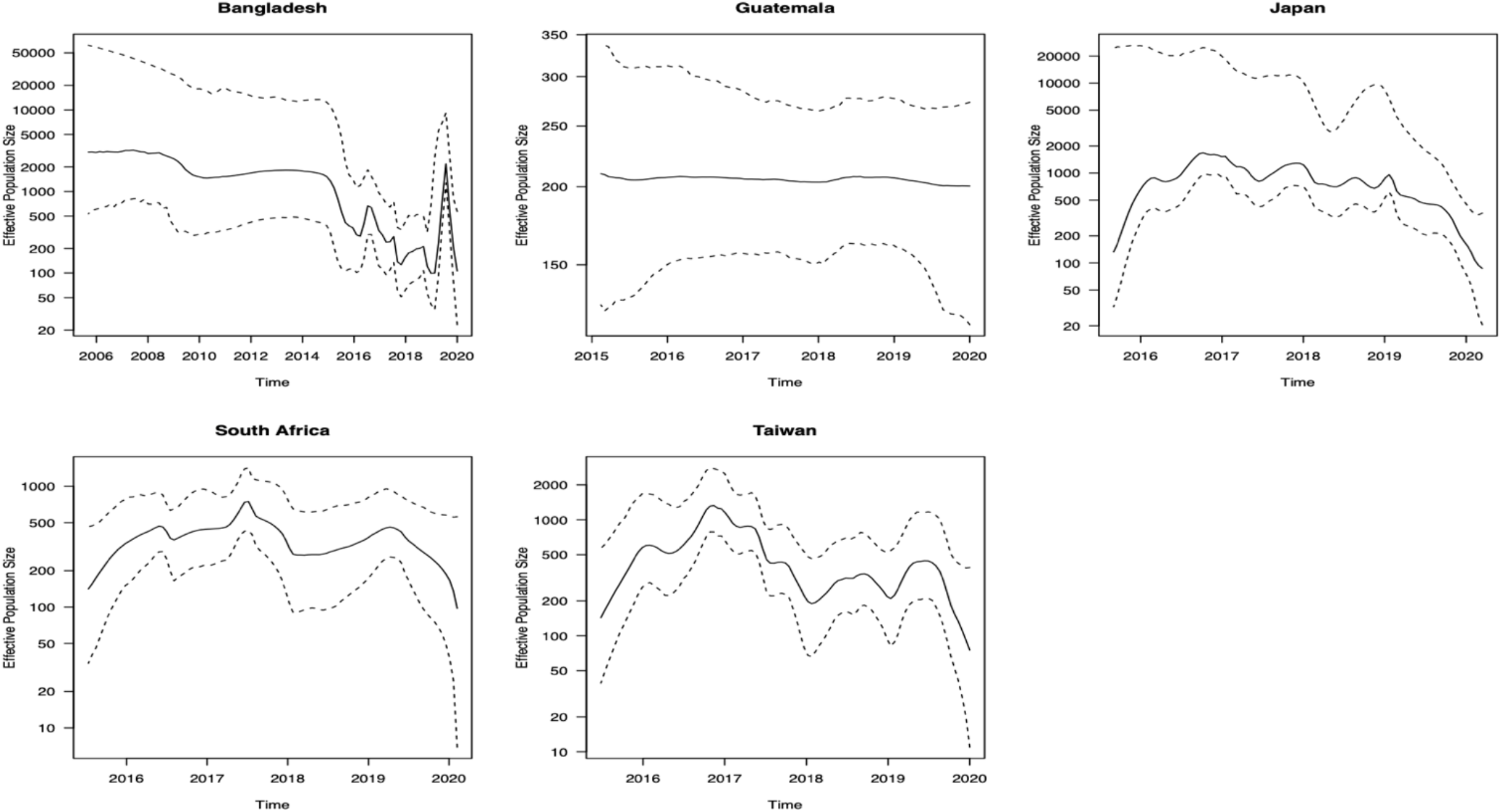
Comparison of Effective Population Size Trajectory for H3N2 in selected regions.

In addition, we would like to thank the following institutions for providing the sequences obtained from GISAID:

Yokohama City Institute of Public Health., National Institute of Animal Health, China Center For Disease Control And Prevention, Akita Research Center for Public Health and Environment, Ningbo international travel healthcare center, University of Corsica, Tottori Prefectural Institute of Public Health and Environmental Science, Ishikawa Prefectural Institute of Public Health and Environmental Science, Zhejiang Provincial Center for Disease Control and Prevention, Research Institute for Environmental Sciences and Public Health of Iwate Prefecture, Nara Prefectural Institute for Hygiene and Environment, Royal Darwin Hospital, Kawasaki City Institute of Public Health, Wuhan Institute of Virology, Tochigi Prefectural Institute of Public Health and Environmental Science, National Institute for Biological Standards and Control (NIBSC), Kyoto City Institute of Health and Environmental Sciences, UOC Policlinico di Bari DIMO, California Department of Health Services, Laboratorio Aziendale di Microbiologia e Virologia, Cabrini Pathology, Laboratorio Central do Estado do Rio de Janeiro - LACEN, Okinawa Prefectural Institute of Health and Environment, Yamaguchi Prefectural Institute of Public Health and Environment, First Affiliated Hospital Medical School of Zhejiang University, New York State Department of Health, Hubei Provincial center for disease control and prevention, Shimane Prefectural Institute of Public Health and Environmental Science, Yamanashi Institute for Public Health, Beijing Center for Disease Prevention and Control, Kumamoto Prefectural Institute of Public Health and Environmental Science, National Institute of Biological Standards and Control, Dorevtich Pathology, Fondazione IRCCS Policlinico San Matteo, Shiga Prefectural Institute of Public Health, Austin Health, National Institute of Infectious Diseases (NIID), Instituto Adolfo Lutz, State Key Laboratory of Virology and Wuhan Institute of Virology, Chinese Academy of Sciences, Royal Melbourne Hospital, Fukui Prefectural Institute of Public Health, Kobe Institute of Health, Institute of Medical and Veterinary Science (IMVS), Prince of Wales Hospital, Laboratorio Nacional De Salud Guatemala, Melbourne Pathology, Hyogo Prefectural Institute of Public Health and Consumer Sciences, Osaka City Institute of Public Health and Environmental Sciences, Saitama Institute of Public Health, Universita degli Studi di Palermo, Dipartimento di Scienze per la Promozione della Salute, Ehime Prefecture Institute of Public Health and Environmental Science, Wakayama Prefectural Research Center of Environment and Public Health, Aichi Prefectural Institute of Public Health, WHO Collaborating Centre for Reference and Research on Influenza, Kagawa Prefectural Research Institute for Environmental Sciences and Public Health, Fukushima Prefectural Institute of Public Health, Sapporo City Institute of Public Health, Amedeo di Savoia Hospital, LACEN, AOU Meyer, Instituto Oswaldo Cruz FIOCRUZ - Laboratory of Respiratory Viruses and Measles (LVRS), Nagasaki Prefectural Institute for Environment Research and Public Health, University of Sassari, Tokushima Prefectural Centre for Public Health and Environmental Sciences, John Hunter Hospital, Friedrich-Loeffler-Institut, New York City Department of Health, Instituto de Salud Publica de Chile, WHO Chinese National Influenza Center, The Kyodoken Institute for Animal Science Research, Universita Cattolica del Sacro Cuore, Centers for Disease Control and Prevention, University of Florence, National Institute for Medical Research, Childrens Hospital Westmead, Saitama City Institute of Health Science and Research, Alfred Hospital, University of Trieste, Chiba Prefectural Institute of Public Health, Okayama Prefectural Institute for Environmental Science and Public Health, Laboratorio di Virologia - Azienda Ospedaliero-Universitaria Ospedali Riuniti - Ancona, Miyazaki Prefectural Institute for Public Health and Environment, Osaka Prefectural Institute of Public Health, Niigata City Institute of Public Health and Environment, Fukuoka Institute of Public Health and Environmental Sciences, Hiroshima City Institute of Public Health, University of Padova, Kochi Public Health and Sanitation Institute, Utsunomiya City Institute of Public Health and Environment Science, Beijing Mentougou District Center for Diseases Prevention and Control, FUNED MG, Hobart Pathology, University of Siena, Virus Research Center, Sendai Medical Center, Istituto Superiore di Sanita, Ningbo International Travel Healthcare Center, University of Milan, Kanagawa Prefectural Institute of Public Health, San Diego County Public Health Lab, Kyoto Prefectural Institute of Public Health and Environment, Ibaraki Prefectural Institute of Public Health, Nagano Environmental Conservation Research Institute, Gunma Prefectural Institute of Public Health and Environmental Sciences, Shanghai International Travel Healthcare Center, Saitama medical university, Chiba City Institute of Health and Environment, Istituto Superiore di Sanita (ISS), Niigata University (IDRC), CNR Virus des Infections Respiratoires - France SUD, Microbiology Services Colindale, Public Health England, The First Affiliated Hospital, College of Medicine, Zhejiang University, Niigata University (DPH), Sakai City Institute of Public Health, Tokyo Metropolitan Institute of Public Health, Institut Pasteur, Universita di Corsica Pasquale Paoli, Center for Public Health and Environment, Hiroshima Prefectural Technology Research Institute, Canberra Hospital, Clinical Virology Unit, CDIM, Seqirus Pty Ltd (CSL Group), National Institute for Communicable Diseases, WHO Centre for Reference, Beijing Institute of Microbiology and Epidemiology, Institut Pasteur of Shanghai, Victorian Infectious Diseases Reference Laboratory, University of Perugia, Mentougou District center for diseases prevention and control, Oita Prefectural Institute of Health and Environment, National Institute for Communicable Disease, The University of Hong Kong, Institut Pasteur of Shanghai, CAS, Laboratorio Central do Estado do Parana - LACEN, Queensland Health Forensic and Scientific Services, Shizuoka City Institute of Environmental Sciences and Public Health, Kumamoto City Environmental Research Center, Royal Chidrens Hospital, Amagasaki City Institute of Public Health, University of Genoa, Hubei Provincial Center for Disease Control and Prevention, CSL Ltd, icddr,b International Centre for Diarrhoeal Disease Research, Bangladesh, Laboratorio Central de Saude Publica Professor Goncalo Moniz, LACEN-BA, Evandro Chagas Institute, Laboratorio Central de Saude Publica (LACEN RJ), Kagoshima University, Fukuoka City Institute for Hygiene and the Environment, Guangzhou No.8 People, Lismore Base Hospital, Pathwest QE II Medical Centre, State Key Laboratory of Reipiratory Disease (Guangzhou Medical University), Sagamihara City Laboratory of Public Health, University of Parma, South China Agricultural University, Crick Worldwide Influenza Centre, Saga Prefectural Institute of Public Health and Pharmaceutical Research, Yamagata Prefectural Institute of Public Health, LACEN SC, Influenza Surveillance Centre for Disease Control, National Health Laboratory, Saitama Medical University, Kagoshima Prefectural Institute for Environmental Research and Public Health, Microbiology University Politecnica delle Marche, Mie Prefecture Health and Environment Research Institute, Institute of Epidemiology Disease Control and Research (IEDCR), Sendai City Institute of Public Health, Gifu Prefectural Institute of Health and Environmental Sciences, Guangdong Provincial Center for Disease Control and Prevention, Douglass Hanly Moir Pathology, Nagano City Health Center, Westmead Hospital, Shizuoka Institute of Environment and Hygiene, National Influenza Center, Yokosuka Institute of Public Health, Guangzhou Institute of Respiratory Diseases (GIRD), Royal Hobart Hospital, Miyagi Prefectural Institute of Public Health and Environment, Niigata Prefectural Institute of Public Health and Environmental Sciences, Hamamatsu City Health Environment Research Center, Aomori Prefectural Institute of Public Health and Environment, University of Study of Siena, South China Agricultural University Veterinary Medicine College, AO dei Colli Monaldi - Cotugno, U.O.C. Microbiologia e Virologia, Laboratorio Central de Saude Publica de Alagoas, LACEN-AL, Monash Medical Centre, Canterbury Health Services, Wakayama City Institute of Public Health, University of Bari, Universita degli Studi di Padova, Vanderbilt University Medical Center, U.S. Air Force School of Aerospace Medicine, Hokkaido Institute of Public Health, Kitakyusyu City Institute of Enviromental Sciences, Gifu Municipal Institute of Public Health, New York Medical College, and Toyama Institute of Health.

GISAID accession numbers for all sequences used can be found at https://github.com/leonkt/accession-numbers/.

## Notes

### Competing Interest Statement

The authors have declared no competing interest.

